# Describing COVID-19 Patients During The First Two Months of Paxlovid (Nirmatrelvir/Ritonavir) Initiation in a Large HMO in Israel

**DOI:** 10.1101/2022.05.02.22274586

**Authors:** Lilac Tene, Gabriel Chodick, Noga Fallach, Wajeeha Ansari, Tal Distelman-Menachem, Yasmin Maor

## Abstract

**Title:** Describing COVID-19 Patients During The First Two Months of Paxlovid (Nirmatrelvir/Ritonavir) Initiation in a Large HMO in Israel

**Objective:** The objective of this feasibility study was to assess the number of patients that could be included in a future Real World Evidence study, which would be designed to explore the impact of Paxlovid (nirmatrelvir/ritonavir) on patient outcomes and healthcare resource utilization (HCRU). We also intend to assess the comparability of the patients who were treated with Paxlovid versus patients who did not receive the treatment, either because they declined any COVID-19 treatment or were diagnosed with COVID-19 prior to Paxlovid availability.

**Methods:** This retrospective observational secondary data study used data from the Maccabi Healthcare Services database during the identification period of June 1, 2021, to February 28, 2022. The study population included patients with at least one positive SARS-CoV-2 RT-PCR test, or a formal rapid antigen test for SARS-CoV-2, during the identification period, the date of which also served as the COVID-19 diagnosis date. We then divided the study population into the following cohorts: Pre-Paxlovid Time Period and Paxlovid Time Period, which was further split into Paxlovid Treated and Paxlovid Untreated.

**Results:** Application of inclusion and exclusion criteria to the study population rendered 20,284 patients in the Pre-Paxlovid Time Period cohort and 5,542 in the Paxlovid Time Period cohort that were eligible to receive Paxlovid. This resulted in 3,714 in the Paxlovid Treated and 1,810 in the Paxlovid Untreated cohorts.

**Conclusions:** This RWE feasibility study of patients with a positive test for COVID-19 between June 1, 2021 to February 28, 2022 illustrates potential comparability between cohorts, as described by their demographics and characteristics.

## Background & Feasibility Objectives

The COVID-19 pandemic, caused by SARS-CoV-2, resulted in more than 6.1 million deaths worldwide as of April 4, 2022.^1^ SARS-CoV-2 may lead to severe acute respiratory syndrome, resulting in hospitalizations, intensive care unit (ICU) stays, and receipt of invasive mechanical ventilation (IMV). Patients at high risk for progression to severe disease (i.e., hospitalization) include unvaccinated individuals, adults ≥⍰65⍰years, current smokers, and those with certain underlying medical conditions including renal disease, chronic lung disease, cardiovascular disease, neurological disorders, and diabetes.^2^ Until recently, treatment options for COVID-19 patients were limited, and most were given in a hospital setting. Treatments in the early phase of the disease relied primarily on convalescent plasma and monoclonal and bi-clonal antibodies administered by injections or intravenously.^3,4^ In particular, no orally available drugs were available to prevent the deterioration of high-risk patients.

On December 22, 2021, the US Food and Drug Administration (FDA) issued an emergency use authorization (EUA) for a new COVID-19 antiviral, PAXLOVID™ (nirmatrelvir [PF-07321332]/ritonavir). Nirmatrelvir is an orally administered antiviral agent targeting the SARS-CoV-2 3-chymotrypsin–like cysteine protease enzyme (Mpro) essential for viral replication.^5^ Nirmatrelvir is metabolized mainly by CYP3A4, thus co-administration with ritonavir, a CYP3A4 inhibitor, enhances nirmatrelvir pharmacokinetics.^6,7^ EPIC-HR (Evaluation of Protease Inhibition for COVID-19 in High-Risk Patients) was a randomized, double-blind study of 2,246 non-hospitalized, symptomatic, unvaccinated adult patients with COVID-19, who were at high risk of progressing to severe illness. The trial analysis showed an 89% reduction in risk of COVID-19-related hospitalization or death from any cause compared to placebo in patients treated within three days of symptom onset. A similar relative risk reduction of 88% was obtained in a subgroup analysis including patients receiving treatment within five days of symptom onset. Paxlovid also demonstrated a tolerable safety profile.^8^

Maccabi Healthcare Services (MHS) is the second largest health maintenance organization (HMO) in Israel, caring for more than 2.5 million people. Paxlovid became available to eligible MHS members on January 2, 2022. The effectiveness of Paxlovid in terms of morbidity, mortality, and its impact on healthcare resource utilization (HCRU) has yet to be assessed using real world evidence (RWE).

The objective of this feasibility study was to assess the number of patients that could be included in a future RWE study, which would be designed to explore the impact of Paxlovid on patient outcomes and healthcare resource utilization (HCRU). We also intend to assess the comparability of the patients who were treated with Paxlovid in MHS versus patients who did not receive the treatment, either because they declined any COVID-19 treatment or were diagnosed with COVID-19 prior to Paxlovid availability.

## Methods

### Study Design

This retrospective observational secondary data study used data from the MHS database and we identified patients during the period of June 1, 2021, to February 28, 2022 (N=2,540,691), as described in **Figure 1**.

**Figure 1.**
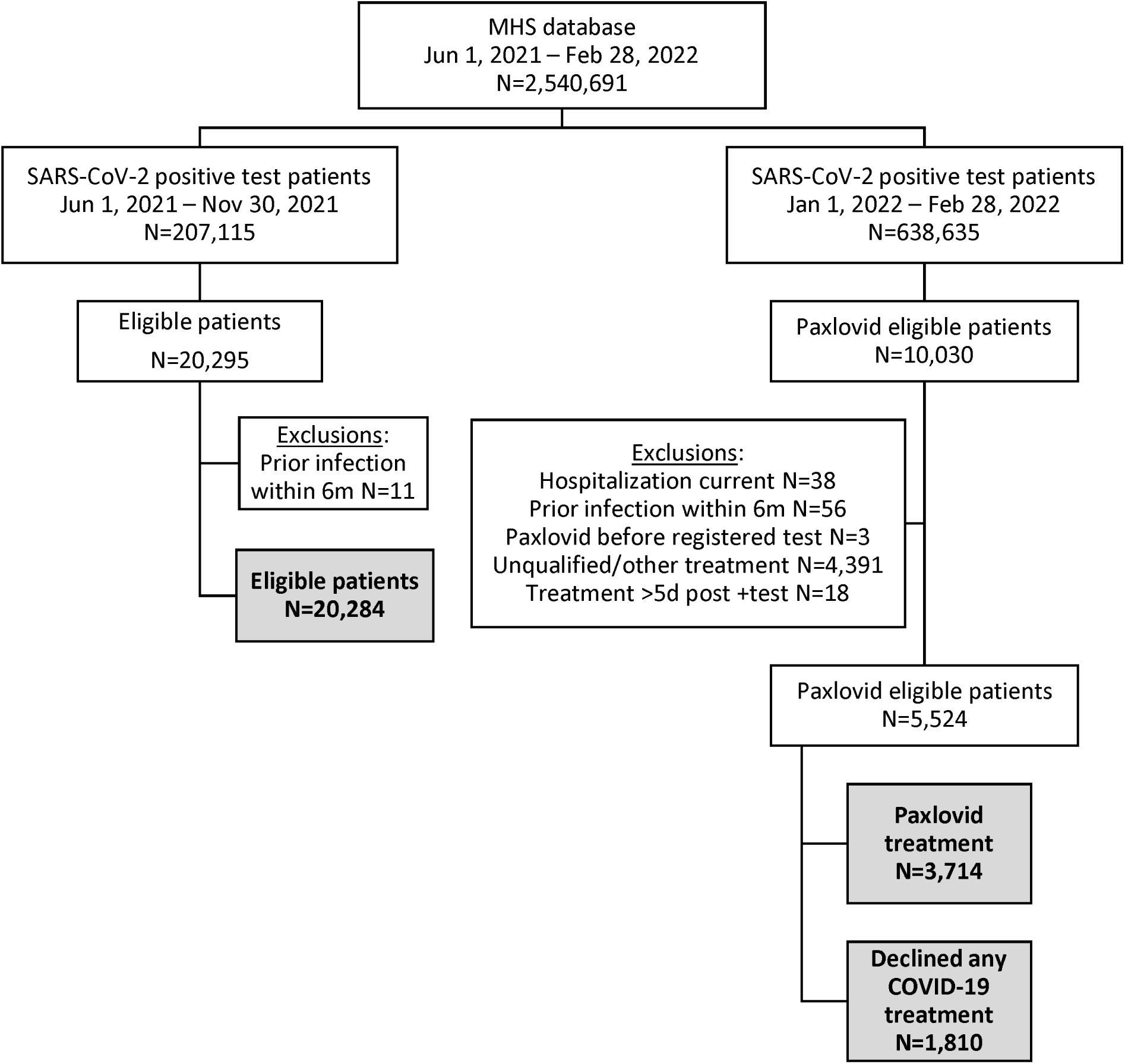
Study flowchart.

MHS is a nationwide health plan (payer-provider) representing a quarter of the population in Israel. The MHS database contains longitudinal data on a stable population of >2.5 million people since 1993 (with <1%/year moving out). Computerized registries of several chronic diseases were developed by MHS and are continuously updated. As a state-mandated health provider and insurer, MHS covers by law all hospitalizations in general hospitals outside the MHS network, as well as visits to urgent care center and ER in hospitals (including cases where a co-pay is required).

Patients’ data are retrieved from MHS medical electronic files and include sociodemographic details, co-morbidities according to ICD-9 codes, immunocompromised registry eligibility, dates of all SARS-CoV-2 PCR tests and formal antigen tests, concomitant medications, hospitalization, and death. COVID-19 symptoms were collected from MHS patients via a questionnaire.

### Study Population

To identify the study population, we included only patients with at least one positive SARS-CoV-2 RT-PCR test, or a formal rapid antigen test for SARS-CoV-2, during the identification period (N=845,750), the date of which also served as the COVID-19 diagnosis date. We divided these patients into the following cohorts:

1. **Pre-Paxlovid Time Period**: Patients in the period when Paxlovid was not available in Israel and Delta was the dominant circulating variant (June 1, 2021, to November 30, 2021, N=207,115)^9^
2. **Paxlovid Time Period**: Patients in the time period when Paxlovid was available in Israel (January 2, 2022, to February 28, 2022, N=638,635), which was then further subdivided into:
  a. **Paxlovid Treated**: patients with a SARS-CoV-2 infection who received Paxlovid treatment
  b. **Paxlovid Untreated**: patients with a SARS-CoV-2 infection who were offered Paxlovid and declined Paxlovid or any other COVID-19 treatment

For patients in the **Paxlovid Treated** cohort, the index date was defined as the date of first dispensed Paxlovid. For patients in the **Paxlovid Untreated** cohort, the index date was defined as the date that Paxlovid treatment was declined.

For patients in the **Pre-Paxlovid Time** Period, the index date was defined using the average number of days from the first positive SARS-CoV-2 test to Paxlovid dispensed date in the **Paxlovid Treated** cohort. The number of days obtained from this calculation was added to the date of the **Pre-Paxlovid Time Period** patients’ first SARS-CoV-2 RT-PCR, or formal antigen positive test.

### Eligibility

To be eligible to receive Paxlovid in Israel, patients have to be aged 70 years or older regardless of risk score (as defined below); aged 50 to 69 years and have a risk score of 2 points or more; or aged 12-49 years with a risk score of four points or more.

Patients were excluded if they were hospitalized prior to Paxlovid treatment or on the index date, received an additional positive PCR or formal rapid antigen test for SARS-CoV-2 in the six months prior to the index date, received Paxlovid prior to a registered SARS-CoV-2 positive test, were found ineligible to receive Paxlovid, received any other approved COVID-19 treatment, or received Paxlovid more than five days after diagnosis.

### Risk Score Calculation

MHS utilizes a COVID-19 Risk Score point system that combines age and characteristics for a numeric score ranging from 0 to 4, where 4 represents highest risk of progression to severe COVID-19. To be eligible to receive Paxlovid patients had to be aged 70 years or more regardless of risk score; aged 50 to 69 years and have a risk score of 2 points or more; or aged 12-49 years with a risk score of four points or more. Each of the following characteristics contributed one point to the risk score: BMI ≥ 30 kg/m2; diabetes mellitus; cardiovascular disease (CVD); chronic kidney disease (CKD); chronic liver disease; neurologic disease; active malignancy or malignancy treated during the past 5 years; Immunosuppression (See **Figure 2**); hospitalization in the past 3 years (not including delivery/birth).

**Figure 2.**
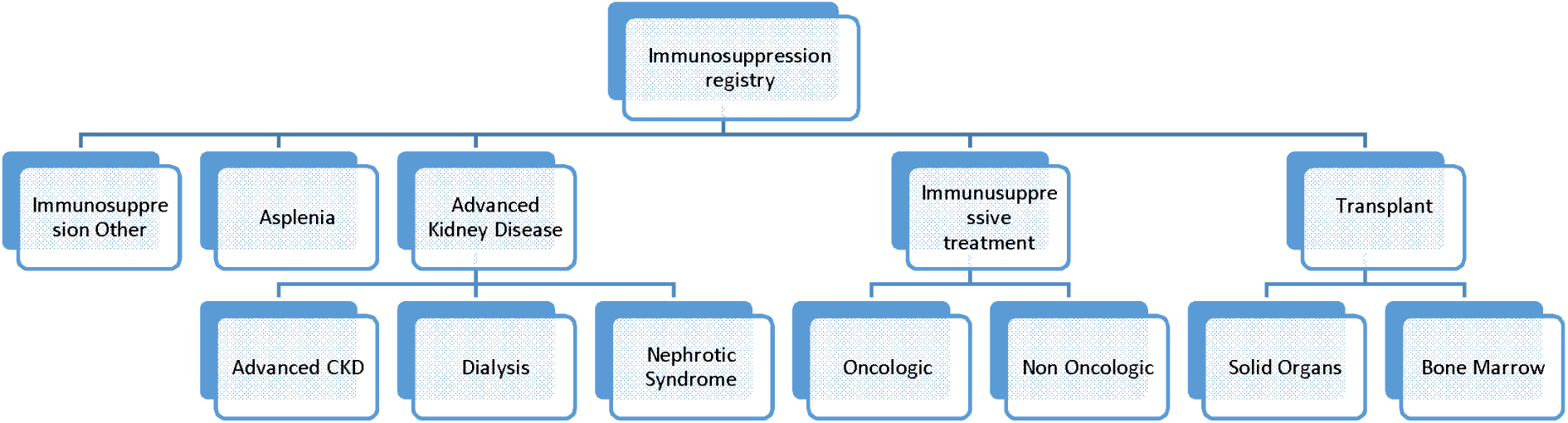
Criteria for inclusion in the MHS immunosuppression registry.

### Statistical Analyses

In this descriptive feasibility study, we explored patients’ data at index date to see if it would be suitable for further analysis. We described continuous variables using summary statistics such as mean, median standard deviation and interquartile range. We also examined lowest and highest observations of each variable to ensure that the data were within plausible ranges. For categorical and binary variables, we calculated counts (missing values and non-missing) and percentages. We calculated the standardized differences (SD) for the continuous and categorical variables.^10^ SD greater than .10 in absolute value suggested potentially meaningful differences. As the purposes of this study were description and feasibility for future study, no formal hypothesis testing was performed. All data analysis was performed by Maccabi Healthcare Services using Statistical Software – SAS v9.4.

## Results

### Study Population Size

In the study population, we identified patients, aged 12 years or older on the index date, that were eligible to receive Paxlovid, as defined by age and their risk score. This rendered 20,284 patients in the **Pre-Paxlovid Time** Period and 10,030 in the **Paxlovid Time Period** that were eligible to receive Paxlovid (**Figure 1**).

To further refine the patients included in the **Paxlovid Time Period** cohort, we applied our exclusion criteria, which resulted in 5,542 patients. Of these, we also excluded an additional 18 patients because they received Paxlovid more than five days after diagnosis.

We then divided patients in the **Paxlovid Time Period** into **Paxlovid Treated** (N=3,714) and **Paxlovid Untreated** (N=1,810) cohorts.

### Characteristics of Study Population

**Table 1** describes the demographics and clinical characteristics of the study population as well as their associated standardize differences.

**Table 1.**
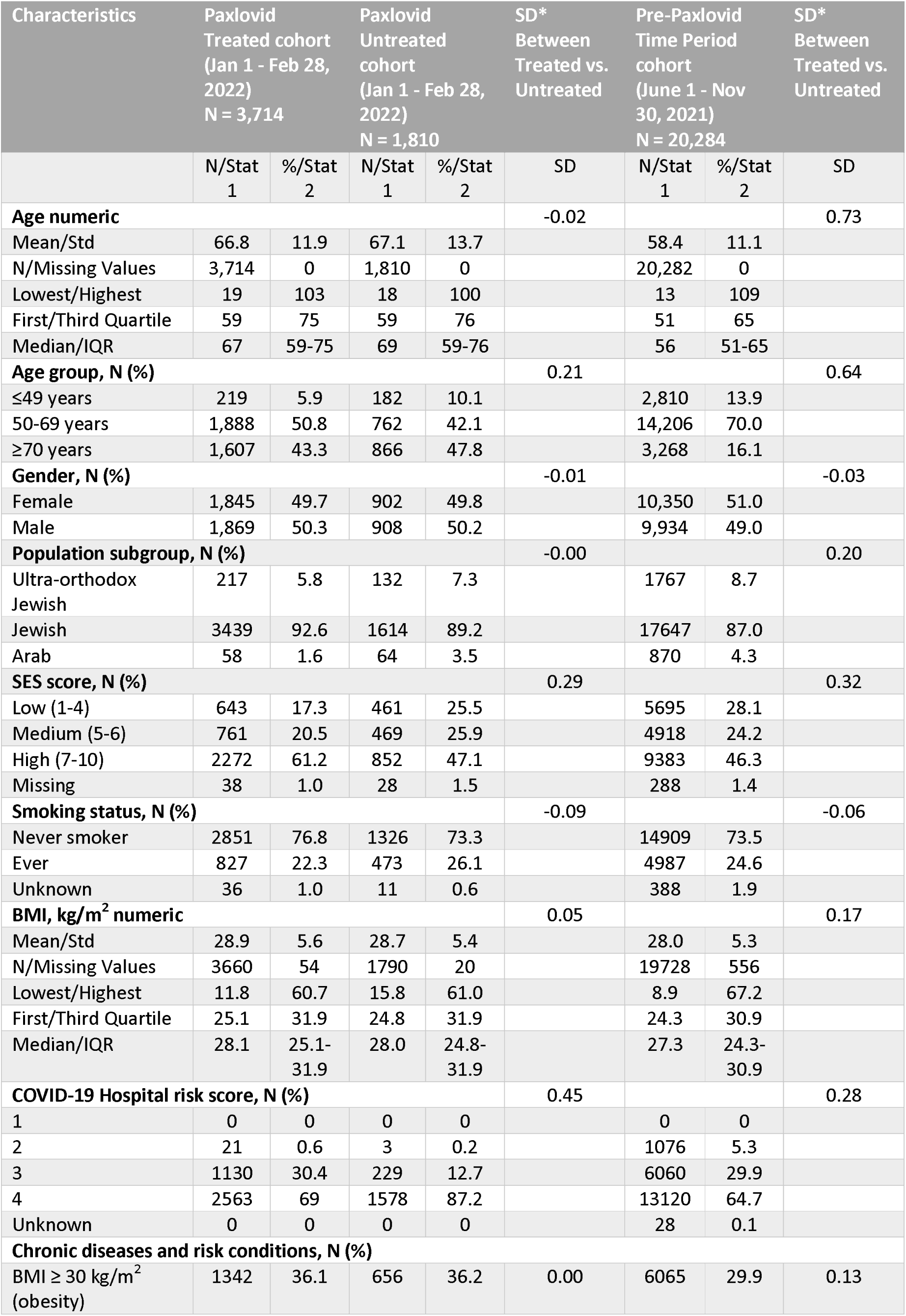
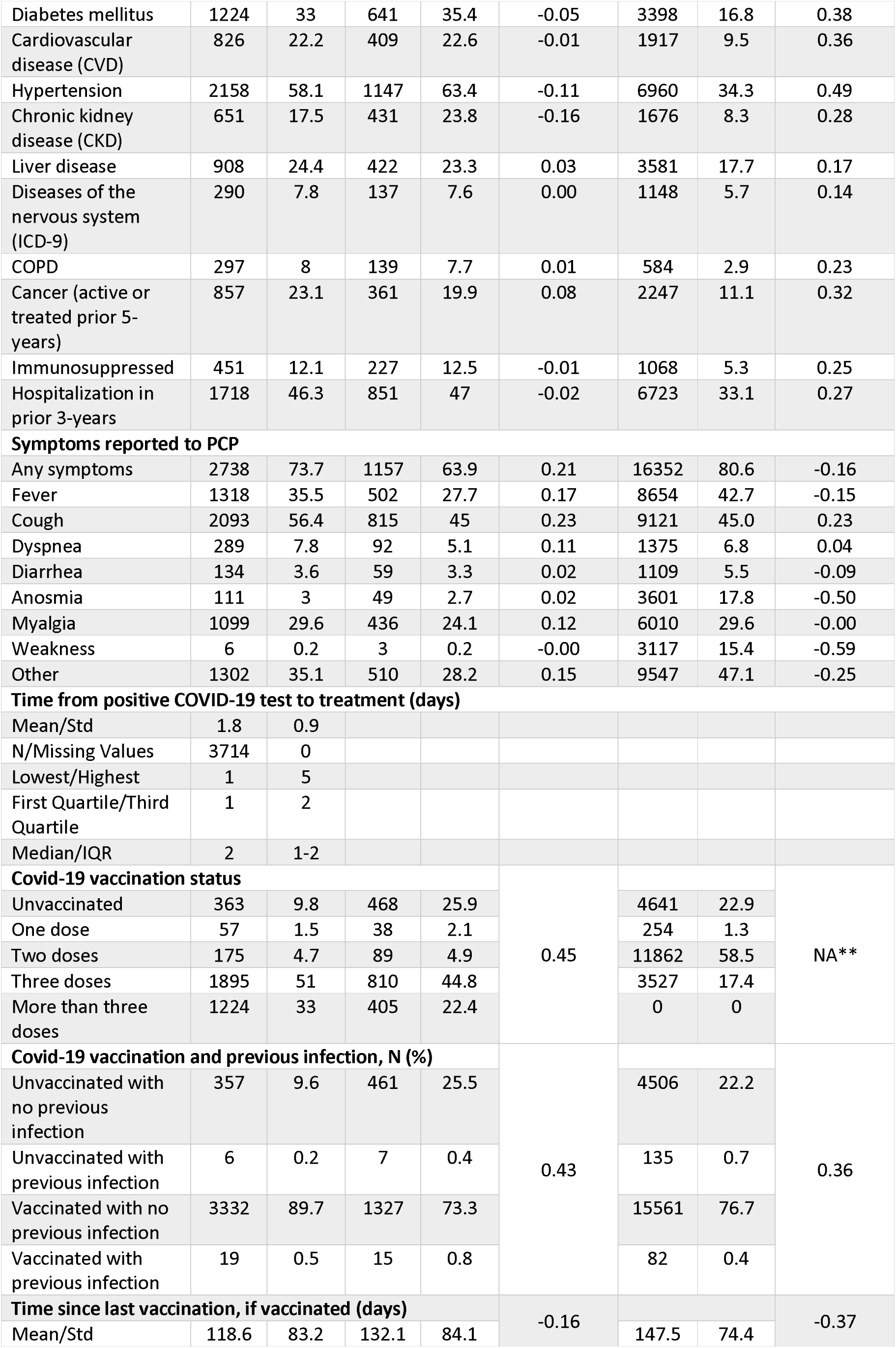
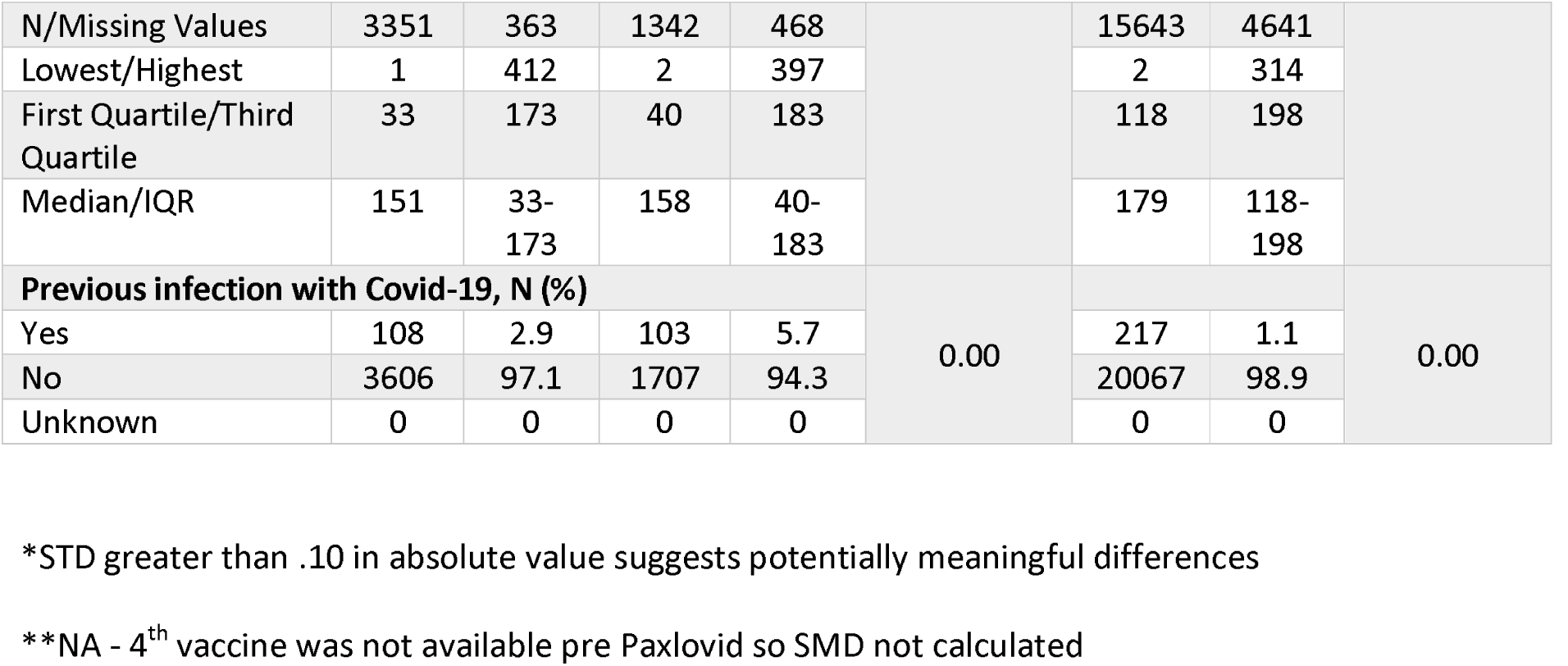
Demographics and Clinical Characteristics.

The proportion of unvaccinated patients was lowest in the **Paxlovid Treated** cohort (9.8%) and similar in the **Paxlovid Untreated** (25.9%) and **Pre-Paxlovid Time** Period (22.9%) cohorts. Most patients had no record of previous COVID-19 infections. The mean time from positive COVID-19 test to treatment was 1.8 days.

The number of patients with a COVID-19 Hospital Risk Score of 4 was highest in the **Paxlovid Untreated** cohort at 87.2%, followed by 69% in the **Paxlovid Treated** and 64.7% in the **Pre-Paxlovid Time Period** cohorts. While some risk conditions varied among the cohorts (e.g., hypertension and CKD being higher in the **Paxlovid Untreated** vs. **Paxlovid Treated**), most were similar, with the **Pre-Paxlovid Time Period** cohort having the lowest frequency of risk conditions.

Most patients reported experiencing at least one COVID-19 symptom across all three cohorts (**Treated**: 74%, **Untreated**: 64%, and **Pre-Paxlovid**: 81%). Of note, fever and cough were less common in the Paxlovid Untreated cohort (27.7% and 45%) versus the **Paxlovid Treated** (35.5% and 56.4%) and the **Pre-Paxlovid Time Period** cohort (80.6% and 45%). On the other hand, anosmia was more common in the **Pre-Paxlovid Time Period** cohort (17.8%) compared to the **Paxlovid Treated** (3%) and Paxlovid Untreated (2.7%) cohorts.

**Paxlovid Treated** patients had higher (7-10) SES scores (61.2%) compared to **Paxlovid Untreated** (47.1%).

In terms of age, the **Paxlovid Time Period** cohorts were generally older than the **Pre-Paxlovid Time Period** cohort (66.8 vs. 58.4 on average). The percentage of patients in the > 70 years age group were higher in the **Paxlovid Treated** (66.8 and 43.3%) and **Paxlovid Untreated** (67.1 and 47.8%) cohorts compared to the **Pre-Paxlovid Time Period** cohort (58.4 and 16.1%). Characteristics associated with an increased risk were similar across cohorts for smoking status and BMI.

## Discussion

The patient population authorized for treatment in Israel differs from the patient population described in the study by Hammond et al.^8^ Paxlovid was authorized for use in Israel for vaccinated and unvaccinated patients, while the trial only included unvaccinated patients. In addition, the Omicron variant was dominant at the time when Paxlovid became available in Israel, while Delta was the dominant variant when the study by Hammond et al. was performed.^8^ These and other differences in the patient population, timing of treatment, and exposure to SARS-CoV-2 in past and present may impact the real world experience of Paxlovid treatment.

This RWE feasibility study of patients with a positive test for COVID-19 between June 1, 2021 to February 28, 2022 illustrates potential comparability between cohorts, as described by their demographics and characteristics.

When comparing the **Paxlovid Treated** cohort to the **Paxlovid Untreated** cohort, the **Paxlovid Treated** cohort had a higher socioeconomic status score, and a lower risk score for progression to severe COVID-19. They were also more likely to be vaccinated and had a shorter interval from the index date and previous vaccine dose. The two groups were similar in their associated comorbidities and number of previous hospitalizations.

Among the three cohorts, the **Paxlovid Treated** and the **Paxlovid Time Period** cohorts were the most different. Patients in the **Pre-Paxlovid Time** Period cohort were younger, had a lower SES score, had a lower BMI, and fewer comorbidities. They also had a lower vaccination rate and a longer time between the last vaccine dose and the index date. These differences may be a result of different health seeking behaviors between the two groups as possibly represented by vaccine acceptance.

Data regarding Paxlovid Time Period patients is still scarce and only few preliminary reports were published comparing patients diagnosed with Omicron and Delta variants. A retrospective cohort study of a large, geographically diverse database of patient electronic health records in the US demonstrated that the monthly incidence rate of COVID-19 infections was 0.5-0.7 when Delta dominated, and rapidly increased to 3.8-5.2 when Omicron dominated.^11^ After propensity-score matching, the risks for severe clinical outcomes in the Omicron cohort were significantly lower than in the Delta cohort. This study did not consider the vaccination status of the study population. Another study assessing patients with COVID-19 in the US demonstrated that a higher proportion of adults admitted during Omicron were fully vaccinated (39.6% versus 25.1%), and fewer received COVID-19-directed therapies when compared to the Delta-dominant period. Fewer fully vaccinated Omicron-dominant period patients died while hospitalized (3.4%) compared with Delta-dominant period patients (10.6%).^12^ The likelihood of ICU admission and death were lowest among adults who had received a booster vaccine dose. Thus the differences seen among the three cohorts in our study are expected and not unique to our cohort.

While it appears a control group for a future RWE study could be identified, additional exclusions or statistical adjustments may be needed to compare patient outcomes. This feasibility supports future analyses by identifying key characteristics needed for future comparisons.

## Data Availability

All data produced in the present work are contained in the manuscript

## Acknowledgements

The authors thank Joanna Atkinson, Griffith Bell, Michael Benigno, Birol Emir, Amie Scott, of Pfizer Inc, for their support in the study design and their contributions to this paper. The authors also thank Arnon Shahar and Shirley Shapiro Ben-David, of Maccabi Healthcare Services, for their contributions to this paper. This study was sponsored by Pfizer Inc. Medical writing support was provided by Yasmin Maor, MD of Tel Aviv University and was funded by Maccabi Healthcare Services.

